# Pharmacokinetics, subject satisfaction, and tolerability of a novel oral iron formulation based on naturally occurring microalgae *Spirulina platensis*

**DOI:** 10.64898/2026.09.15.26363044

**Authors:** Milko Radicioni, Riccardo Assandri, Angelica Bastianello, Emanuela Gentile, Elena P. Tiberio, Federica Sala, Pietro Magrone

## Abstract

**Introduction:** Study to evaluate the pharmacokinetics (PK), subject satisfaction, and gastrointestinal (GI) tolerability of single and multiple doses of a novel oral iron formulation based on iron enriched microalgae *Spirulina platensis* supplemented with inactivated yeast.

**Methods:** Twelve healthy women with baseline serum iron 4–12 µmol/L (22.3–67.0 µg/dL) were admitted to the study site for 3 days in this single centre, multiple-dose, open-label study. Subjects received an oral dose of Monurelle^®^ Ferro Naturale (MFN) on Days 1, 2, and 3. Blood for PK analysis was collected pre-dose (0) and at 1, 2, 3, 4, 5, 8 hours after dosing on Days 1 and 3, with assessment of subject satisfaction and GI tolerability performed 3 hours after each dose.

**Results:** Both single and multiple dosing with MFN led to a defined peak level (C_max_) and increased systemic exposure (AUC_0-t_), with serum iron rising to almost double the circulating amount at 5 h post-dose (t_max_). Mean (±SD) percent change from baseline at 5 h post-dose was 94.7% (±56.1) on Day 1 and 72.0% (±70.7) on Day 3.

The majority of subjects reported being satisfied with the product and would use it again in the future. MFN was very well tolerated, with only one adverse event (AE) reported (episode of vomiting after the first dose, that lasted for one minute and did not recur after the two further doses of MFN). No other subject experienced any GI side effects.

**Discussion:** This first report of oral Monurelle^®^ Ferro Naturale in humans shows this novel formulation to provide a favourable PK profile with respect to serum iron concentrations, with excellent tolerability and high subject satisfaction.

## Introduction

Iron deficiency remains a major nutritional deficiency, affecting an estimated 30% of the global population [Hamarsha et al. 2025]. First-line treatment is generally oral iron supplementation, as it is inexpensive and widely available. However, treatment of iron deficiency with oral iron generally requires daily dosing for extended periods and may be poorly tolerated due to side effects. This can lead to low adherence to therapy, with the leading cause of discontinuation of oral iron being gastrointestinal (GI) side effects, including unpleasant metallic taste, nausea, bloating, constipation, abdominal pain, and diarrhoea [Tolkien et al. 2015]. In addition, many oral iron supplements have low bioavailability, with only 10-20% absorbed and excess iron in the GI tract contributing to poor tolerability [Pantopoulos 2024]. New formulations of iron that have higher bioavailability and improved GI tolerance are needed to improve adherence to oral iron supplementation treatment for iron deficiency [Cancelo-Hidalgo et al. 2013].

A novel formulation of oral iron supplement, Monurelle^®^ Ferro Naturale (MFN) has been developed to optimize gastrointestinal iron absorption and reduce GI disturbance. MFN contains an innovative nutraceutical formulation, registered as Ironatural^®^, consisting of homogeneous mechanical blended iron-enriched *Spirulina* (*Arthrospira platensis* RCC6351) biomass and thermally inactivated yeast (*Saccharomyces cerevisiae* var. *boulardii* SGSb01 cells) at a weight ratio of 9:1 (w/w) [Piccolo et al. 2026]. *Spirulina platensis* is a naturally occurring nutrient-rich blue-green microalgae that grows in warm alkaline lakes in tropical and sub-tropical areas. *Spirulina platensis* is included in the US Food and Drug Administration (FDA) ‘Generally Recognized As Safe’ (GRAS) database and is already used as a dietary supplement, as it is rich in proteins, essential fatty acids, vitamins, minerals and bioactive compounds [Khan et al. 2005].

A number of studies of *Spirulina* have shown promising results with respect to serum iron and haematological parameters. One study showed *Spirulina* supplementation for 12 weeks to increase mean corpuscular haemoglobin in both male and female subjects aged ≥50 years [Selmi et al. 2011]. Eight weeks of 1 g/day *Spirulina* supplementation in adults with ulcerative colitis significantly increased serum iron and improved anaemia parameters compared to placebo [Moradi et al. 2023]. Moderately malnourished children who received 3 g *Spirulina* showed increased serum ferritin and iron levels [Abed et al. 2016].

*Spirulina platensis* present in MFN is cultivated in closed photobioreactors under controlled environmental conditions. To maximize intracellular iron accumulation while preserving productivity, a two-step biofortification strategy is employed: standard cultivation to the stationary phase is followed by biomass harvesting and subsequent resuspension in an aqueous solution enriched with soluble iron salts and metal–organic complexes. The resulting biofortified biomass is collected, thermally dried (< 40°C) to a residual moisture content below 10%, and homogeneously blended with the inactivated yeast component [Piccolo et al. 2026].

The iron-enriched *Spirulina* is supplemented with inactivated yeast (*S. cerevisiae var. boulardii*). The inclusion of this component is intended to support a formulation designed to promote physiologically favourable intestinal conditions for iron regulation and assimilation. Previous preclinical studies have described biological activities associated with inactivated yeast preparations and yeast-derived components (often referred to as postbiotic preparations), including effects on intestinal barrier integrity, gut homeostasis and modulation of inflammatory pathways [Qin et al 2024; Jin Y et al. 2024, Xu et al. 2023].

In a nonclinical study MFN showed iron content comparable to a commercially available sucrosomial iron formulation (MFN 19.68 mg Fe/g of formulation; sucrosomial iron formulation 22.37 mg Fe/g) [Piccolo et al. 2026]. and exhibited lower toxicity on viability of intestinal cells compared to the comparators. Furthermore, MFN showed higher bioaccessibility following simulated GI digestion (96.0%) compared with sucrosomial iron (56.0%), and higher transepithelial iron permeation after 1 hour (90.7% vs 82.9%). At the molecular level MFN resulted in upregulation of key factors involved in iron uptake (divalent metal transporter 1), iron storage (ferritin heavy chain 1) and iron export (ferroportin) [Piccolo et al. 2026].

The primary objective of the current study was to investigate pharmacokinetics (PK), subject satisfaction, and GI tolerability following single and multiple oral administrations of MFN to healthy women.

## Materials and Methods

This study is registered in the UK’s Clinical Study Registry (ISRCTN78497162).

### Study Subjects

Eligible subjects were healthy females aged 18-60 years with serum iron 4.0 – 12.0 µmol/L (22.3 – 67.0 µg/dL) inclusive. Subjects were required to have body mass index (BMI) 18.5-30 kg/m^2^; normal vital signs and ECG; no history of significant disease or allergies that could affect the outcome of the study; negative pregnancy test; and to be in days 5-20 of the menstrual cycle during the interventional phase of the study. All medication (including over-the-counter medication, herbal remedies and nutritional supplements [including iron supplements]) was stopped at least 2 weeks prior to the study. Hormonal contraception was allowed with all subjects required to use a reliable method of contraception. Subjects were excluded from participation if they suffered from metrorrhagia; had donated blood within 3 months of study start; had an abnormal diet (<1600 or >3500 kcal/day) within 4 weeks of the study; were vegetarian, pregnant or lactating; had a positive drug or alcohol test at baseline; or history of drug abuse or consumption of excess alcohol, coffee or tobacco in previous 12 months.

All subjects were required to give written informed consent to participate in the study. The protocol and informed consent form were approved by the local Independent Ethics Committee (Canton of Ticino, Switzerland), and the study was performed in accordance with the Declaration of Helsinki, and Good Clinical Practice (ICH-GCP).

### Study Design

The study was a single centre, multiple-dose, single-way, open-label design and was performed at a Phase I Unit in Switzerland (Cross Research S.A.). Subjects were screened for eligibility to participate in the study within 15 days before first dose (Day 1) and were admitted to the study site on the evening of Day -1. During admission, standardized meals were served with avoidance of iron rich foods. One cup of coffee or tea was allowed after each meal only, with foods containing xanthines (cola, chocolate etc.), and alcohol and grapefruit prohibited. One cigarette (or e-cigarette with nicotine) was allowed after each meal only, with e-cigarettes without nicotine prohibited.

On Day 1 subjects received one capsule of MFN in the morning and blood samples for PK analysis were collected pre-dose (0) and at 1, 2, 3, 4, 5, 8 hours post-dose. A product intake global evaluation and subject satisfaction questionnaire was completed at 3 hours post-dose. Gastrointestinal tolerability was assessed at 3 hours post-dose for any symptoms of heartburn, stomach discomfort / pain, abdominal discomfort / pain, nausea, vomiting (using a 3-point scale). On Day 2 and Day 3 subjects received a second and third dose of MFN respectively with the same post-dose assessments performed on Day 3 as for Day 1. Subjects were discharged from the Phase I unit on Day 3 after collection of the 8-hour post-dose blood sample and after physical examination, vital signs check and laboratory analysis (haematology, blood chemistry including iron balance, and urinalysis).

Blood samples for PK analysis were collected using an indwelling catheter with switch valve. The cannula was rinsed after each sampling with 1 mL of sterile saline solution. The first 1 mL of blood was discarded at each collection point.

### Study Medication

Study medication in the form of Monurelle^®^ Ferro Naturale (MFN) was provided by Zambon S.p.A., (manufacturer Genelife S.r.l.) as capsules for oral administration containing 500 mg of *Spirulina platensis* of which 30 mg is iron (batch 250738; expiry date December 2028). The dose of iron 30 mg is the maximum daily amount of iron recommended as a food supplement in Italy [Marangoni et al. 2025] and is below the safe level of intake established by the European Food Safety Authority [EFSA 2024]. Study medication was stored at room temperature (15-25°C). One capsule of MFN was administered to each subject under fasting condition once daily at 08:00±1 h for 3 consecutive days. MFN capsules were to be swallowed (without chewing) with 150 mL still mineral water at room temperature.

### Measurement of serum iron concentration

Concentration of iron in serum was determined at a central laboratory (Unilabs Ticino, Switzerland) according to ISO 15189:2022. Samples were kept at room temperature at the study centre for at least 30 min and for a maximum of 60 min and then centrifuged at 2500×g for 10 min at room temperature. Serum samples were then stored in pre-labelled polypropylene tubes at 2-8°C until shipment at the end of Days 1 and 3 to the central laboratory for analysis, with a second back-up sample stored at Cross Research at -20°C.

### Statistical Analysis

The primary endpoints were serum iron concentrations and PK profile and parameters, with secondary endpoints of subject evaluation of product intake and satisfaction. Safety and tolerability endpoints included the frequency of adverse events (AEs) occurring or worsening after first dose of study medication, GI symptoms identified on GI tolerability evaluations based on a 3-point scale (reported as AEs), physical examination, vital signs (BP, heart rate), body weight, and laboratory parameters.

The sample size was not based on any formal sample size calculation but estimated to be sufficient for the descriptive purposes of the study.

PK parameters were presented using descriptive statistics (including geometric mean) for quantitative variables and frequencies for qualitative variables. Pharmacokinetic analyses were performed on both observed (non-baseline-corrected) and baseline-corrected serum iron concentrations. For each subject, baseline was calculated as the arithmetic mean of the serum iron concentrations measured at -12 h on Day -1 and pre-dose on Day 1; this value was subtracted from each post-dose concentration, and negative baseline-corrected values were set to zero. Pharmacokinetic parameters were derived using Phoenix WinNonlin^TM^ version 8.3.5 and SAS^®^ version 9.3 (TS1M1) and 9.4 (M9) after single (Day 1) and multiple (Day 3) dose administration of study medication. Demographic and safety data were analysed using SAS^®^ version 9.3 (TS1M1) and 9.4 (M9).

Missing data was not replaced, with all non-available data evaluated as missing values.

## Results

### Study Subjects

Of the 46 subjects screened, 12 subjects met the inclusion and exclusion criteria and were enrolled between 17 February 2026 and 27 February 2026. There were no important findings with respect to demographics, baseline characteristics, medical history or concomitant medication. Median age was 49 years (range 24-59), and median BMI was 23.8 (range 20.7 to 29.9). Eight of the 12 enrolled subjects were of reproductive age, with the remaining 4 subjects at least 1-year post-menopause. At study entry no subjects were taking any medication or herbal remedy, except for one subject who was receiving oral contraceptives. There were no significant findings at baseline for vital signs, ECG, or laboratory results. Individual baseline serum iron concentration ranged from 25.7 to 92.2 μg/dL.

All subjects received the planned study medication and completed the study, with no protocol violations. One subject was excluded from the PK analysis due to an episode of vomiting occurring on Day 1 after administration of study medication.

### Pharmacokinetics

Mean (± SD) serum concentrations of iron after single (Day 1) and multiple (Day 3) doses of MFN are shown in Table 1 and Figure 1. Baseline-corrected concentrations followed a similar pattern and are reported here in text. On Day 1 iron concentrations exceeded the baseline level at all time points for all subjects, with the same result observed after multiple dosing (Day 3) in 7 out of 11 subjects.

**Figure 1:**
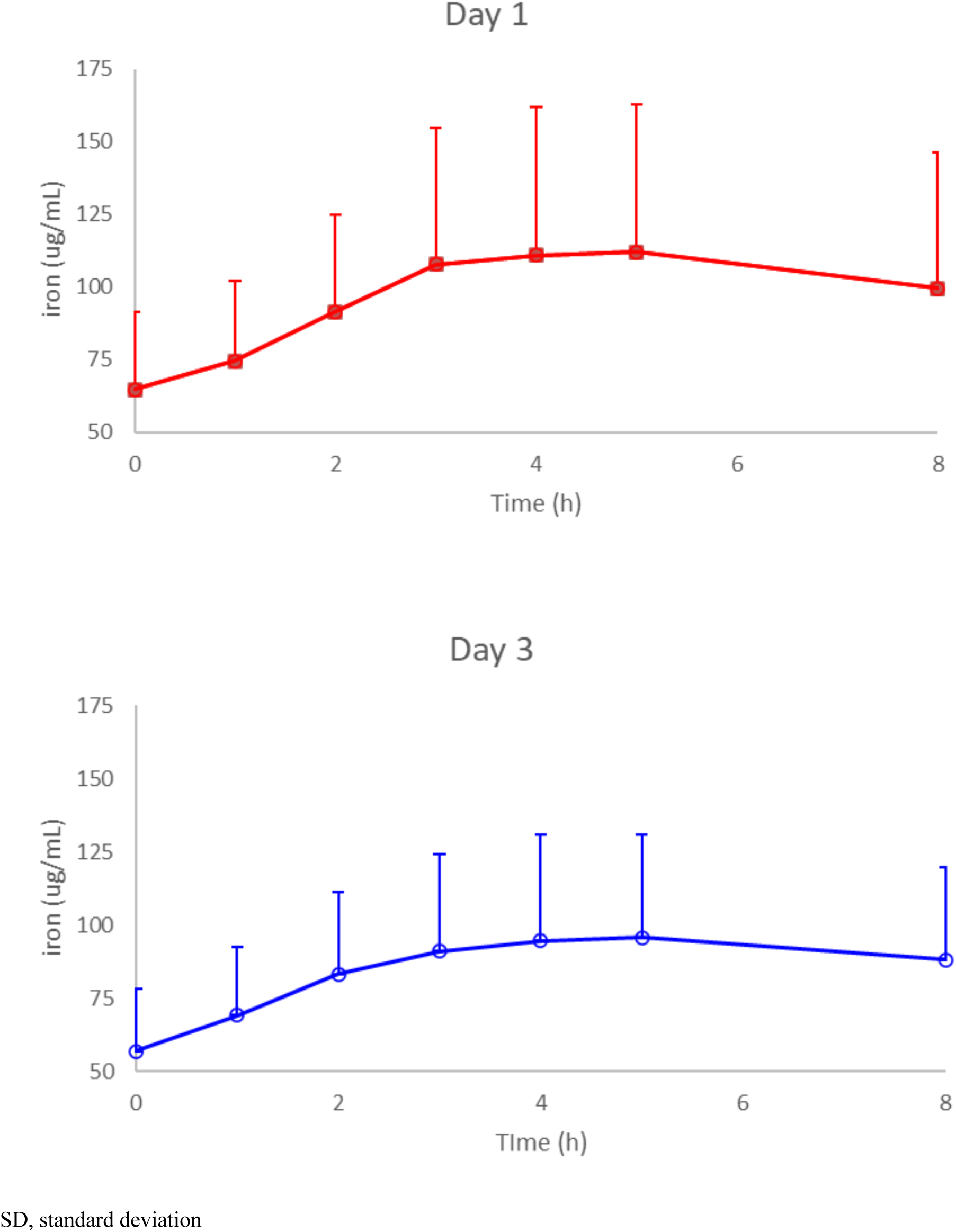
Observed (non-baseline corrected) serum iron concentrations over time after single (Day 1) and multiple (Day 3) doses of MFN (mean [SD], n=11).

**Table 1:** Serum iron concentration (µg/dL) after single (Day 1) and multiple (Day 3) doses of study medication (mean [±SD], n=11)

| **Time** | **Day 1 (single dose)** | **Day 3 (multiple dose)** |
| --- | --- | --- |
| **-12 h pre-dose** | 53.15 ± 21.25 | - |
| **Pre-dose** | 64.68 ± 26.47 | 56.85 ± 21.15 |
| **1 h** | 74.64 ± 27.49 | 69.21 ± 23.47 |
| **2 h** | 91.53 ± 33.32 | 83.11 ± 27.96 |
| **3 h** | 107.67 ± 47.10 | 91.03 ± 32.92 |
| **4 h** | 110.94 ± 50.74 | 94.65 ± 36.13 |
| **5 h** | 111.94 ± 50.97 | 95.74 ± 34.90 |
| **8 h** | 99.49 ± 46.74 | 88.07 ± 31.69 |

SD, standard deviation

Iron concentrations were generally higher on Day 1 than on Day 3 at all time-points; at 5 h post-dose, mean (±SD) observed serum iron concentrations were 111.94 ± 50.97 µg/dL on Day 1 and 95.74 ± 34.90 µg/dL on Day 3, while the corresponding baseline-corrected concentrations were 53.02 ± 38.26 µg/dL and 37.12 ± 28.51 µg/dL, respectively. Mean (±SD) percent change from baseline at 5 h post-dose was 94.7% (±56.1) on Day 1 and 72.0% (±70.7) on Day 3.

Following both single and multiple doses of MFN the concentration-time profiles were comparable, with iron levels rising to 5 h post-dose and slightly decreasing thereafter (Figure 1). An absorption curve was appreciable in most individual subjects on both Day 1 and Day 3, whereas in general no clear elimination curve could be defined.

A defined peak level (C_max_) was observed following both single and multiple product administrations, with slightly higher values on Day 1 compared to Day 3 (Table 2). The corresponding mean (±SD) baseline-corrected C_max_ values were 55.45 ± 37.51 µg/dL on Day 1 and 39.92 ± 27.92 µg/dL on Day 3. Observed AUC_0-t_ values are reported in Table 2; the corresponding mean (±SD) baseline-corrected AUC_0-t_ values were 316.00 ± 215.20 h·µg/dL on Day 1 and 227.22 ± 182.74 h·µg/dL on Day 3. Median time to peak (t_max_) was 5 h after both single and multiple dosing for observed and baseline-corrected data. At 8 h post-dose, all subjects on Day 1 and 9 out of 11 subjects on Day 3 still showed iron levels above baseline. PK parameters λ_z_, t_1/2_, AUC_0-∞_, and %AUC_extra_ could not be calculated.

**Table 2:** Serum iron PK parameters after single (Day 1) and multiple (Day 3) doses of MFN (n=11)

| PK parameter | Day 1 (single dose) | Day 3 (multiple dose) |
| --- | --- | --- |
| **C<sub>max</sub> (µg/dL), mean (± SD)** | 114.36 ± 50.20 | 98.84 ± 33.76 |
| **AUC<sub>0-t</sub> (h\*µg/dL), mean (± SD)** | 787.34 ± 325.54 | 690.00 ± 243.21 |
| **t<sub>max</sub> (h), median (range)** | 5 (3 – 8) | 5 (2 – 8) |

SD, standard deviation

Inter-subject variability (CV%) was relatively high, ranging from 57.9% to 91.1% on Day 1 and from 69.0% to 147.8% on Day 3.

### Product intake and subject satisfaction evaluations

Table 3 shows the results for product intake and subject satisfaction evaluations. MFN was judged as easy or very easy to swallow by all subjects (100%) at all assessments. Six subjects on Day 1 and two subjects on Days 2 and 3 reported a metallic aftertaste, describing it as mild or very mild.

**Table 3:** Product intake and subject satisfaction evaluation (n=12)

| Question | Answer | n (%) |  |  |
| --- | --- | --- | --- | --- |
|  |  | Day 1 | Day 2 | Day 3 |
| How do you rate the ease of swallowing of the product? | Very easy | 8 (66.7) | 11 (91.7) | 10 (83.3) |
|  | Easy | 4 (33.3) | 1 (8.3) | 2 (16.7) |
|  | Neither easy nor uneasy | 0 (0.0) | 0 (0.0) | 0 (0.0) |
|  | Uneasy | 0 (0.0) | 0 (0.0) | 0 (0.0) |
|  | Very uneasy | 0 (0.0) | 0 (0.0) | 0 (0.0) |
| Did you feel a metallic aftertaste after the product intake? | Yes | 6 (50.0) | 2 (16.7) | 2 (16.7) |
|  | No | 6 (50.0) | 10 (83.3) | 10 (83.3) |
| If YES, how do you rate this aftertaste? | Not applicable | 6 (50.0) | 10 (83.3) | 10 (83.3) |
|  | Very mild | 0 (0.0) | 2 (16.7) | 0 (0.0) |
|  | Mild | 6 (50.0) | 0 (0.0) | 2 (16.7) |
|  | Neither mild nor strong | 0 (0.0) | 0 (0.0) | 0 (0.0) |
|  | Strong | 0 (0.0) | 0 (0.0) | 0 (0.0) |
|  | Very strong | 0 (0.0) | 0 (0.0) | 0 (0.0) |
| How do you rate your satisfaction with the product? | Very satisfying | 5 (41.7) | 7 (58.3) | 7 (58.3) |
|  | Satisfying | 5 (41.7) | 3 (25.0) | 5 (41.7) |
|  | Neither satisfying nor unsatisfying | 2 (16.7) | 2 (16.7) | 0 (0.0) |
|  | Unsatisfying | 0 (0.0) | 0 (0.0) | 0 (0.0) |
|  | Very unsatisfying | 0 (0.0) | 0 (0.0) | 0 (0.0) |
| Would you repeat the use of this product in the future? | Certainly yes | 8 (66.7) | 8 (66.7) | 8 (66.7) |
|  | Probably yes | 3 (25.0) | 4 (33.3) | 3 (25.0) |
|  | Neither yes nor no | 1 (8.3) | 0 (0.0) | 1 (8.3) |
|  | Probably no | 0 (0.0) | 0 (0.0) | 0 (0.0) |
|  | Certainly no | 0 (0.0) | 0 (0.0) | 0 (0.0) |

On Day 1 and Day 2 a total of 10 (83.3%) of subjects reported being satisfied or very satisfied with MFN, with the remaining 2 subjects reported neither satisfying nor unsatisfying. By Day 3 all subjects (100%) reported being satisfied or very satisfied with study medication. All subjects on Day 2 and 91.7% on Day 1 and Day 3 reported that they would repeat the use of MFN in the future.

### Safety and Tolerability

Overall MFN was very well tolerated with no serious AEs or AEs leading to study discontinuation. Only one AE occurred during the study, in a subject who experienced vomiting approximately one hour after intake of study medication on Day 1 that lasted around 1 minute, resolved spontaneously, and did not recur after the two subsequent administrations of study medication. This single episode of vomiting was reported as an AE of moderate intensity and assessed by the investigator to be related to study medication.

No clinically significant changes in laboratory parameters, vital signs, physical examination, or body weight were observed during the study.

#### Gastrointestinal tolerability

GI tolerability of MFN was very good, and apart from the single episode of vomiting on Day 1 reported as an AE, all other subjects experienced no GI symptoms (Table 4).

**Table 4:** Gastrointestinal tolerability evaluations (n=12)

| Question | Answer | n (%) |  |  |
| --- | --- | --- | --- | --- |
|  |  | Day 1 | Day 2 | Day 3 |
| Heartburn | 0 – None | 12 (100) | 12 (100) | 12 (100) |
|  | 1 - Mild | 0 (0.0) | 0 (0.0) | 0 (0.0) |
|  | 2 - Moderate | 0 (0.0) | 0 (0.0) | 0 (0.0) |
|  | 3 - Severe | 0 (0.0) | 0 (0.0) | 0 (0.0) |
| Stomach discomfort | 0 – None | 12 (100) | 12 (100) | 12 (100) |
|  | 1 - Mild | 0 (0.0) | 0 (0.0) | 0 (0.0) |
|  | 2 - Moderate | 0 (0.0) | 0 (0.0) | 0 (0.0) |
|  | 3 - Severe | 0 (0.0) | 0 (0.0) | 0 (0.0) |
| Stomach pain | 0 – None | 12 (100) | 12 (100) | 12 (100) |
|  | 1 - Mild | 0 (0.0) | 0 (0.0) | 0 (0.0) |
|  | 2 - Moderate | 0 (0.0) | 0 (0.0) | 0 (0.0) |
|  | 3 - Severe | 0 (0.0) | 0 (0.0) | 0 (0.0) |
| Abdominal discomfort | 0 – None | 12 (100) | 12 (100) | 12 (100) |
|  | 1 - Mild | 0 (0.0) | 0 (0.0) | 0 (0.0) |
|  | 2 - Moderate | 0 (0.0) | 0 (0.0) | 0 (0.0) |
|  | 3 - Severe | 0 (0.0) | 0 (0.0) | 0 (0.0) |
| Abdominal pain | 0 – None | 12 (100) | 12 (100) | 12 (100) |
|  | 1 - Mild | 0 (0.0) | 0 (0.0) | 0 (0.0) |
|  | 2 - Moderate | 0 (0.0) | 0 (0.0) | 0 (0.0) |
|  | 3 - Severe | 0 (0.0) | 0 (0.0) | 0 (0.0) |
| Nausea | 0 – None | 12 (100) | 12 (100) | 12 (100) |
|  | 1 - Mild | 0 (0.0) | 0 (0.0) | 0 (0.0) |
|  | 2 - Moderate | 0 (0.0) | 0 (0.0) | 0 (0.0) |
|  | 3 - Severe | 0 (0.0) | 0 (0.0) | 0 (0.0) |
| Vomiting | 0 – None | 11 (91.7) | 12 (100) | 12 (100) |
|  | 1 - Mild | 0 (0.0) | 0 (0.0) | 0 (0.0) |
|  | 2 - Moderate | 1 (8.3) | 0 (0.0) | 0 (0.0) |
|  | 3 - Severe | 0 (0.0) | 0 (0.0) | 0 (0.0) |

## Discussion

This is the first study in humans of a novel formulation of oral iron, a preparation based on the naturally occurring microalgae *Spirulina platensis* enriched with iron and supplemented with inactivated yeast.

The study showed a substantial increase in serum iron following the intake of MFN containing 30 mg of iron. Iron levels increased on average for up to 5 h post-dose with a slight decrease thereafter. At 5 h post-dose, the observed mean concentrations were 111.94 µg/dL on Day 1 and 95.74 µg/dL on Day 3, while the corresponding mean baseline-corrected concentrations were 53.02 µg/dL and 37.12 µg/dL. The mean percent changes from baseline were 94.7% on Day 1 and 72.0% on Day 3. Pre-dose iron concentrations on Day 3 were comparable to baseline, indicating no apparent accumulation with daily dosing over 3 days. The lower post-dose concentrations observed on Day 3 compared with Day 1 are likely within the inherent variability of serum iron measurements and study procedures rather than indicative of a meaningful difference in iron exposure. Both single and multiple dosing of MFN led to a defined peak level (C_max_) and resulted in measurableAUC_0-t_ values, indicating systemic iron absorption following administration. Mean time to peak (t_max_) was 5 h post-dose after both single and multiple administrations. While an absorption curve was observed in most study subjects, an elimination curve could not be defined in any subject. This may be attributed to the fact that C_max_ occurred at 5 h post-dose, while sampling was carried out only up to 8 h after dosing.

MFN demonstrated an excellent tolerability profile, with no subjects withdrawing from the study because of tolerability or safety. A single AE was reported, but it was transient, did not recur after the two subsequent administrations of MFN, and the subject expressed a favourable opinion with respect to product satisfaction and GI tolerability. No other subjects experienced any GI symptoms following administration of MFN over 3 days. Most participants did not perceive any metallic taste following MFN administration on Days 2 and 3 and in those who described a metallic taste, this was assessed as mild or very mild. This observation is of relevance in clinical practice, as the persistence of a metallic taste is one of the factors contributing to poor compliance in iron supplementation treatment [Tolkien et al. 2015].

MFN received very positive subject evaluations, with 83% of subjects reporting being satisfied or very satisfied on Day 1, and this percentage increased to 100% on Day 3. All subjects (100%) on Day 2 and 91.7% on Days 1 and 3 would use the product again in the future.

Although the study provides important information on the novel MFN formulation, the study design has some limitations including the open-label design (although this would not be expected to influence the PK results), a relatively short duration (3 days) with respect to GI tolerability and the absence of a comparator arm. Although there was no comparator in the current study, the PK profile with MFN is comparable to that observed in previously studied iron formulations with similar C_max_ and AUC_0-t_. In a study of 9 healthy women, sucrosomial iron was used as the reference product (formulation also contained 70 mg of vitamin C) [Cupone et al. 2023]. A single dose of sucrosomial iron resulted in a mean serum iron AUC_0-8_ of 837 μg x h/dL and C_max_ of 131.2 μg/dL. Median t_max_ with sucrosomial iron was reported to be 2 hours, considerable shorter than that observed for MFN (5 h) in the current study, suggesting a slower absorption rate of MFN, which may be beneficial with respect to GI tolerability. The test product in the study by Cupone et al. (IBSA orodispersible iron film with 30 mg of iron as pyrophosphate and 400 µg of folic acid) showed a similar PK profile to sucrosomial iron.

In summary, this is the first report of the administration in humans of Monurelle^®^ Ferro Naturale, a novel formulation of oral iron based on the naturally occurring microalgae *Spirulina platensis* enriched with iron and supplemented with inactivated yeast. The study demonstrates a favourable PK profile of the new formulation, consistent with increased iron availability in the systemic circulation, together with excellent tolerability and high subject satisfaction.

## Acknowledgments

The authors are grateful to all study participants who took part in this research. They also thank the clinical and operational teams at CROSS Research S.A. for their support in the conduct of the study. Medical writing assistance in the preparation of this manuscript was provided by Dr. Diana Barkley (Seedhopper GmbH) and funded by Zambon S.p.A. in accordance with Good Publication Practice (GPP3) guidelines

## Author Contributions

All authors have read and approved the manuscript and agree to be accountable for all aspects of the work.

Conceptualization (MR, RA, EG, FS, PM), investigation (MR), methodology (MR, RA, EG, PM, FS, ET), project administration (PM, ET), supervision (MR, PM), writing – original draft, and writing (MR, AB, ET) – review & editing (MR, RA, AB, EG, ET, FS, PM).

## Data Availability

The research data described in this manuscript are not shared.

## Funding

The study was sponsored by Zambon S.p.A.

## Author Disclosures

Emanuela Gentile, Elena Tiberio, Federica Sala and Pietro Magrone are current employees of Zambon S.p.A.

Angelica Bastianello is an employee of Cultipharm S.r.l., developer of the Ironatural^®^ ingredient used in Monurelle^®^ Ferro Naturale.

Milko Radicioni and Riccardo Assandri are employees of CROSS Research S.A., the contract research organization that conducted the study.

The authors declare no other competing interests.

## Notes

### Competing Interest Statement

The authors have declared no competing interest.

### Clinical Trial

ISRCTN78497162

### Author Declarations

Ethics committee of Canton Ticino (Switzerland) gave ethical approval for this work

